# Extremely high SARS-CoV-2 seroprevalence in a strictly-Orthodox Jewish community in the UK

**DOI:** 10.1101/2021.02.01.21250839

**Authors:** Katherine M Gaskell, Marina Johnson, Victoria Gould, Adam Hunt, Neil RH Stone, William Waites, Ben Kasstan, Tracey Chantler, Sham Lal, Chrissy h. Roberts, David Goldblatt, Rosalind M Eggo, Michael Marks

**Affiliations:** Department of Clinical Research, London School of Hygiene & Tropical Medicine, Keppel Street, London. WC1E 7HT UK; Great Ormond Street Institute of Child Health Biomedical Research Centre, University College London; Hospital for Tropical Diseases, University College London Hospital NHS Foundation Trust, London; Centre for Mathematical Modelling of Infectious Diseases, London School of Hygiene & Tropical Medicine, Keppel Street, London. WC1E 7HT UK; Centre for Health, Law and Society, University of Bristol Law School, Bristol. BS1 1RJ; Department of Global Health and Development, London School of Hygiene & Tropical Medicine, Keppel Street, London. WC1E 7HT UK

**Author notes:** **Author for Correspondence:** Dr Michael Marks, Clinical Research Department, Faculty of Infectious and Tropical Diseases, London School of Hygiene & Tropical Medicine, London, WC1E 7HT, United Kingdom.

## Abstract

**Background:** Ethnic and religious minorities have been disproportionately affected by SARS-CoV-2 worldwide. The UK strictly-Orthodox Jewish community has been severely affected by the pandemic. This group shares characteristics with other ethnic minorities including larger family sizes, higher rates of household crowding and relative socioeconomic deprivation. We studied a UK strictly-Orthodox Jewish population to understand how COVID-19 had spread within this community.

**Methods:** We performed a household-focused cross-sectional SARS-CoV-2 serosurvey specific to three antigen targets. Randomly-selected households completed a standardised questionnaire and underwent serological testing with a multiplex assay for SARS-CoV-2 IgG antibodies. We report clinical illness and testing before the serosurvey, seroprevalence stratified by age and gender. We used random-effects models to identify factors associated with infection and antibody titres.

**Findings:** A total of 343 households, consisting of 1,759 individuals, were recruited. Serum was available for 1,242 participants. The overall seroprevalence for SARS-CoV-2 was 64.3% (95% CI 61.6-67.0%). The lowest seroprevalence was 27.6% in children under 5 years and rose to 73.8% in secondary school children and 74% in adults. Antibody titres were higher in symptomatic individuals and declined over time since reported COVID-19 symptoms, with the decline more marked for nucleocapsid titres.

**Interpretation:** In this tight-knit religious minority population in the UK, we report one of the highest SARS-CoV-2 seroprevalence levels in the world to date. In the context of this high force of infection, all age groups experienced a high burden of infection. Actions to reduce the burden of disease in this and other minority populations are urgently required.

**Funding:** This work was jointly funded by UKRI and NIHR [COV0335; MR/V027956/1], a donation from the LSHTM Alumni COVID-19 response fund, HDR UK, the MRC and the Wellcome Trust. The funders had no role in the design, conduct or analysis of the study or the decision to publish. The authors have no financial relationships with any organizations that might have an interest in the submitted work in the previous three years; no other relationships or activities that could appear to have influenced the submitted work.

**Research In Context:** *Evidence before the study:* In January 2020, we searched PubMed for articles on rates of SARS-CoV-2 infection amongst ethnic minority groups and amongst the Jewish population. Search teams included “COVID-19”, “SARS-CoV-2”, seroprevalence, “ethnic minority”, and “Jewish” with no language restrictions. We also searched UK government documents on SARS-CoV-2 infection amongst minority groups. By January 2020, a large number of authors had reported that ethnic minority groups experienced higher numbers of cases and increased hospitalisations due to COVID-19. A small number of articles provided evidence that strictly-Orthodox Jewish populations had experienced a high rate of SARS-CoV-2 infection but extremely limited data was available on overall population level rates of infection amongst specific ethnic minority population groups. There was also extremely limited data on rates of infection amongst young children from ethnic minority groups.

*Added value of the study:* We report findings from a population representative, household survey of SARS-CoV-2 infection amongst a UK strictly Orthodox Jewish population. We demonstrate an extremely high seroprevalence rate of SARS-CoV-2 in this population which is more than five times the estimated seroprevalence nationally and five times the estimated seroprevalence in London. In addition the large number of children in our survey, reflective of the underlying population structure, allows us to demonstrate that in this setting there is a significant burden of disease in all age groups with secondary school aged children having an equivalent seroprevalence to adults.

*Implications of the available evidence:* Our data provide clear evidence of the markedly disproportionate impact of SARS-CoV-2 in minority populations. In this setting infection occurs at high rates across all age groups including pre-school, primary school and secondary school-age children. Contextually appropriate measures to specifically reduce the impact of SARS-CoV-2 amongst minority populations are urgently required.

## Introduction

The UK has been severely affected by the COVID-19 pandemic with 66,197 deaths involving COVID-19 up to the 27th November 2020^1^. In the UK the pandemic has disproportionately affected minority ethnic populations with relatively higher numbers of positive cases and overrepresentation in admissions to hospitals and Intensive Care Units and deaths.^2–4^. Similar disparities have been observed in other settings and amongst hospitalised patients, mortality has been found to be higher in ethnic minority patients in both the UK and USA^5–7^. The reasons for this disparity are unclear but are likely multifactorial, reflecting both higher rates of infection resulting from socio-economic factors including deprivation, less ability to work from home, higher use of public transport and larger household sizes, as well as increased severity of illness due to higher rates of comorbidities or delayed access to care^8^.

Most population based studies in the UK have focused on the impact experienced by larger minority ethnic groups and not on smaller groups, such as the Jewish community, despite comprising approximately 265,000 individuals or 0.5% of the UK’s population^9^. In particular, strictly Orthodox Jewish communities have anecdotally reported high rates of infection, morbidity and hospitalisation during the first wave of the UK pandemic^10^. These anecdotal reports are supported by findings from Public Health England (PHE) which suggests a higher risk from infection in the UK Jewish population and higher rates of death due to COVID-19 in those self identifying as Jewish with an age standardised mortality rates between March and May for Jewish men over 65 years of 759 per 100,000 population, much higher than in other religious groups^11^. This higher rate of infection amongst the Jewish community might reflect socio-demographic differences compared to the general UK population. Strictly Orthodox Jewish communities have among the highest total fertility rates in the UK, and experience high rates of overcrowded homes and higher levels of socio-economic deprivation ^12,13^. However, after adjusting for socio-economic factors the hazard ratio for death in Jewish men remained double that of Christian men suggesting that routine socio-economic factors alone do not explain all of the increased rate of infection seen in this community^11^. During the first wave of the COVID-19 pandemic in Israel, strictly Orthodox Jewish communities also reported higher incidence rates of COVID-19 than other socio-economically similar communities^14,15^, or when compared to households in Arab communities with similar sizes and levels of crowding^15^.

In Spring 2020, a UK tightly-knit strictly-Orthodox Jewish community became aware that they appeared to be experiencing a high burden of SARS-CoV-2. In view of this, and national data suggesting a high burden of infection in the Jewish community and a possible disproportionately high burden in the strictly Orthodox community, we co-developed a cross-sectional serological survey to measure the burden of infection and identify factors associated with transmission and to inform local control efforts.

## Methods

### Recruitment and survey methodology

Within a strictly Orthodox Jewish community in the UK we conducted a household-focused seroprevalence survey between late-October and early December 2020 prior to the third national lockdown. We obtained a comprehensive list of all resident households within the community, held by our community collaborators, and randomly selected households for the study. A second list of households where laboratory confirmed infections were known to have occured (the ‘enriched’ list) was also included to inform subsequent transmission models. Members of the study team telephoned households to complete a standardised questionnaire, including demographics, comorbidities, report of any previous presumed COVID-19 illness, previous PCR or serological testing for SARS-CoV-2, access to care whilst unwell with a COVID-19 like illness, illness severity, attendance at work, educational or other community locations and travel overseas between February - November 2020. All households were visited within 10 days of completing the questionnaire, all participants had blood drawn for a serum sample except where a member of the household reported active symptoms of COVID-19 in which case the visit was scheduled for a minimum of 14 days after symptom onset. All members of the household were eligible to be included without age restrictions.

### Laboratory analysis

Serum samples were analysed for the presence of IgG specific for SARS CoV-2 trimeric spike protein (S), Receptor Binding Domain (RBD) and nucleocapsid (N) antigens using a multiplex chemiluminescence immunoassay (MSD, Rockville, MD) evaluated by our laboratory as previously described ^16,17^.

### Statistical analysis

Assuming 10% seroprevalence and a design effect of 2 we calculated we would need to recruit 1,730 participants. Assuming an average household size of 6 we estimated this would be approximately 300 households.

To obtain unbiased estimates, we restricted our analysis to individuals from randomly selected households. Results related to the ‘enriched’ household dataset will be presented in future analyses. We used cut-off values for the SARS-CoV-2 immunoassay from previous validation studies for antibodies against trimeric-spike, nucleocapsid and receptor binding domain. For the purpose of this analysis, we considered a positive trimeric-spike response as evidence of a prior SARS-CoV-2 infection as this target was shown to be the most sensitive and specific target in assay validation^16^. As assay cut-offs are well defined in adults but less well defined in children, we conducted a sensitivity analysis in which we considered thresholds for the trimeric-spike assay of twice the previously defined limit.

We report the proportion of individuals who had a clinical illness consistent with COVID-19 and the proportion of individuals who had accessed testing prior to this survey. We calculated the seroprevalence of SARS-CoV-2 infection stratified by age and gender and adjusted for clustering at the household level. Age was classified as pre-school (0-4 years), primary education (5-10 years), secondary education (11-18 years), adults and retirement (greater than 67 years). We created a proxy variable which accounted for the number of community events attended during the year (hereinafter ‘community-gatherings’) and a variable reflecting household overcrowding based on the number of household residents and the number of bedrooms^18^. To identify factors associated with SARS-CoV-2 infection we fitted a multivariable random-effects logistic regression model adjusted for clustering at the household level. We hypothesised that age, gender, being unable to work from home, attendance at community events and household overcrowding were *a priori* likely to be associated with an increased risk of infection. We assessed relationships between titres of antibodies against each of SARS-CoV-2 spike, receptor binding domain and nucleocapsid in relation to individuals age, gender, the presence or absence of a symptomatic illness, and for symptomatic individuals the timing of their illness using a log-linear random effects model.

### Ethics

The study was approved by the London School of Hygiene & Tropical Medicine Ethics Committee (*Ref 22532*). Verbal informed consent was given during the telephone survey and written consent provided prior to phlebotomy. Parents provided written consent for children.

## Results

### Enrollment and demographics

A total of 903 randomly selected households were approached, of which 343 households comprising 1,759 individuals were enrolled (Figure 1). An additional 70 households with known cases of COVID-19 were provided by our community partners (referred to as ‘enriched households’) were approached to participate, of which 28 households comprising 183 individuals were enrolled (Supplementary Figure 3). Serum samples were collected from 1242 individuals (70.6%) from 283 randomly selected households and 137 individuals (74.9%) from 24 ‘enriched households.

**Figure 1.**
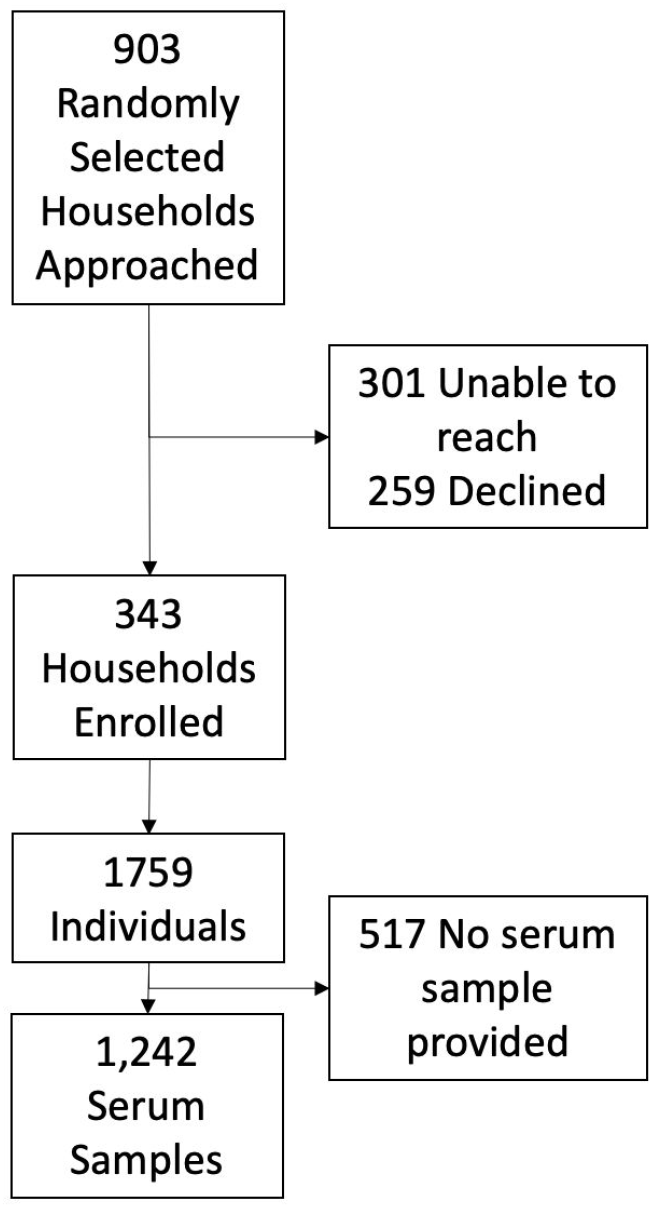
CONSORT diagram for recruitment into the study.

**Figure 2.**
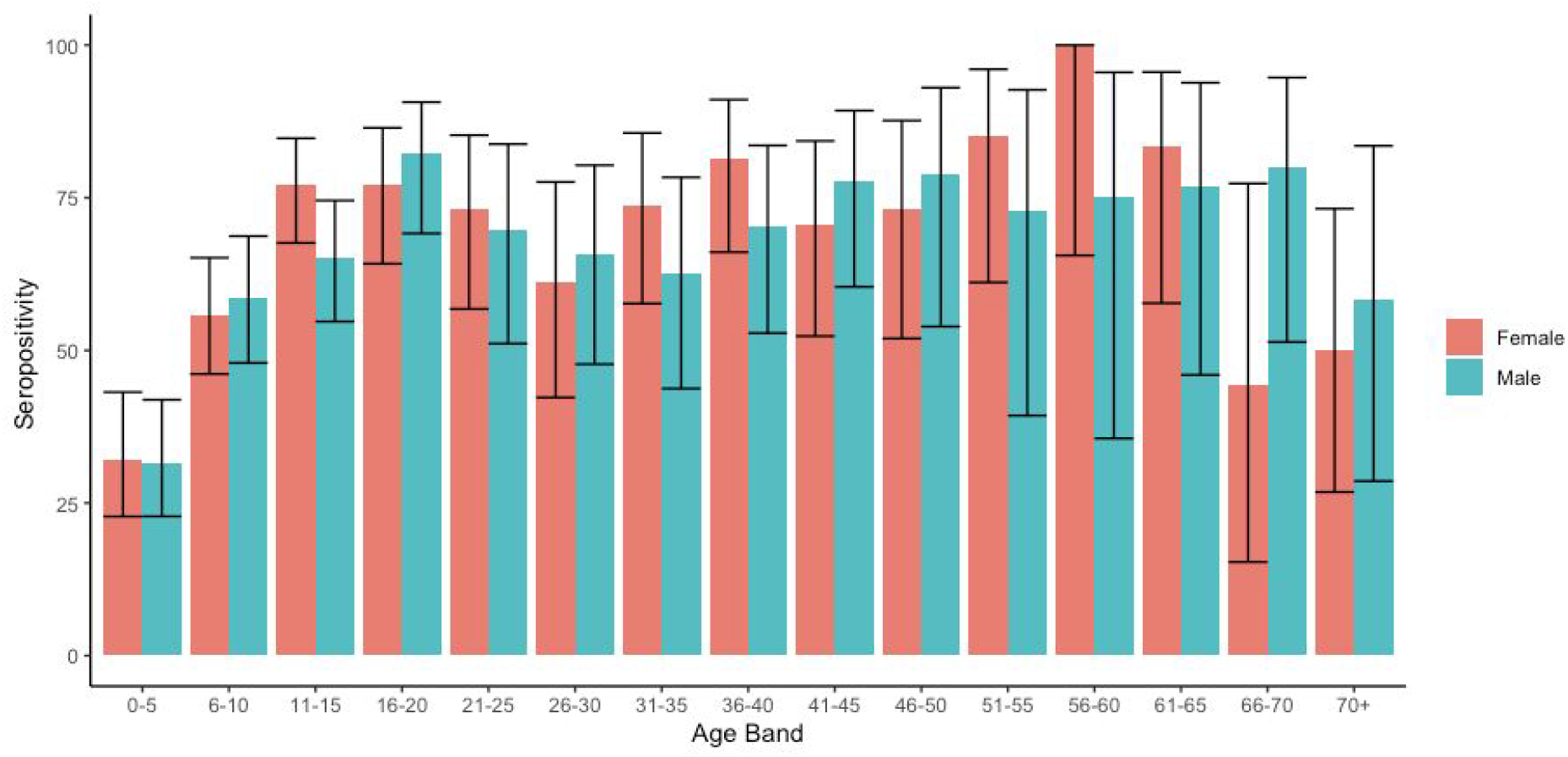
Age specific seroprevalence in participants in the study. Colours indicate male and female participants.

**Figure 3.**
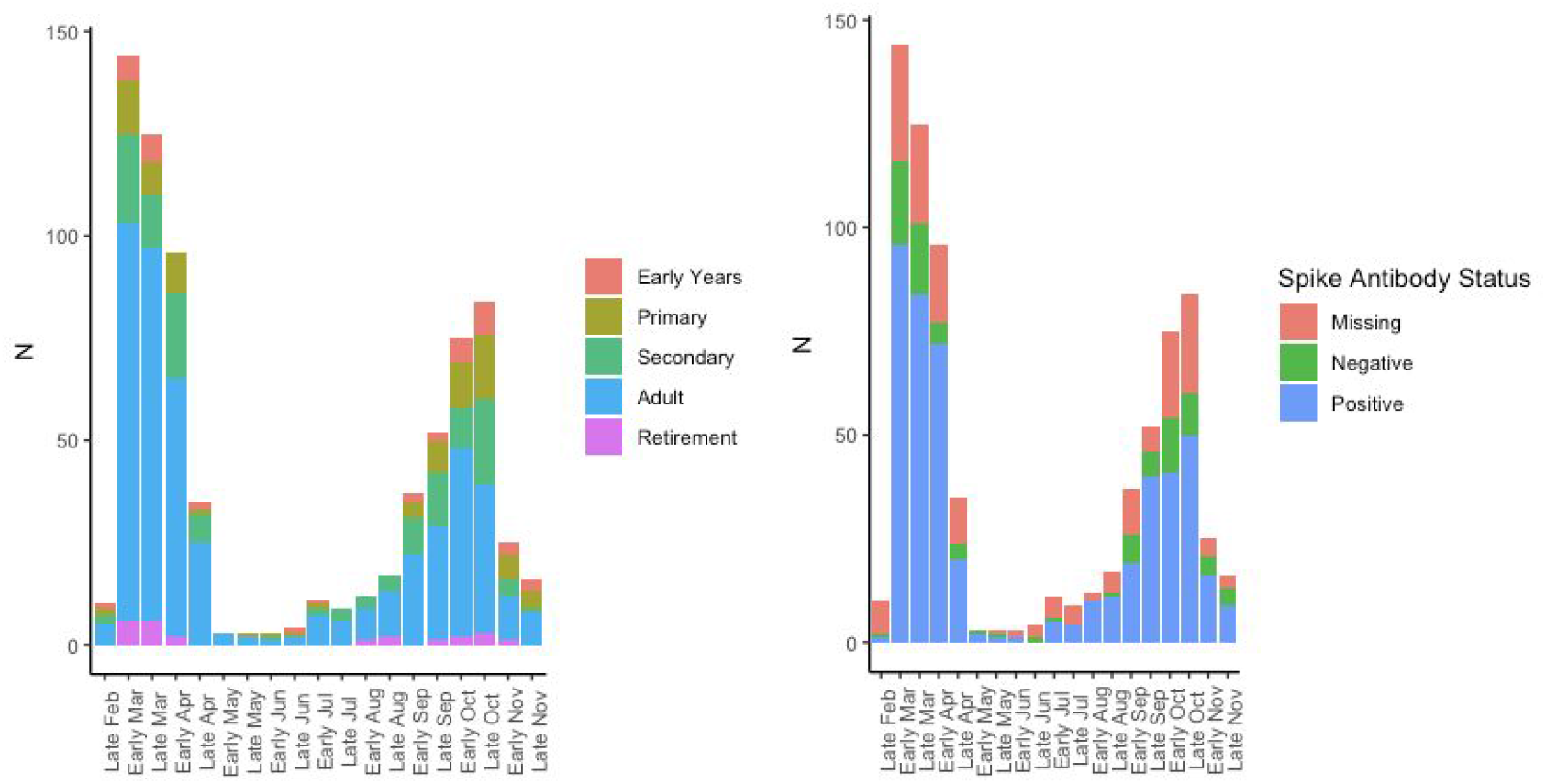
Self-reported COVID-19-like illness in participants in the study stratified by age and antibody status. The reduced number of self-reported cases in November is an artefact reflecting the timing of the survey. Missing Anti-Spike antibody status occured when participants did not participate in phlebotomy after completing the survey. No phlebotomy samples gave an inconclusive result.

The median household size was 5 (IQR 3-7, Range 1-14 cf. UK median 2)^19^ with a median of 3 bedrooms (IQR 2-5). The median age of survey participants was 14 years (IQR 7-33, cd. UK median age 40 years) and 48.6% of participants were male (Supplementary Table 1). Of individuals who gave a serum sample 48% were male and the median age was 16 years (IQR 9-37).

**Table 1.**
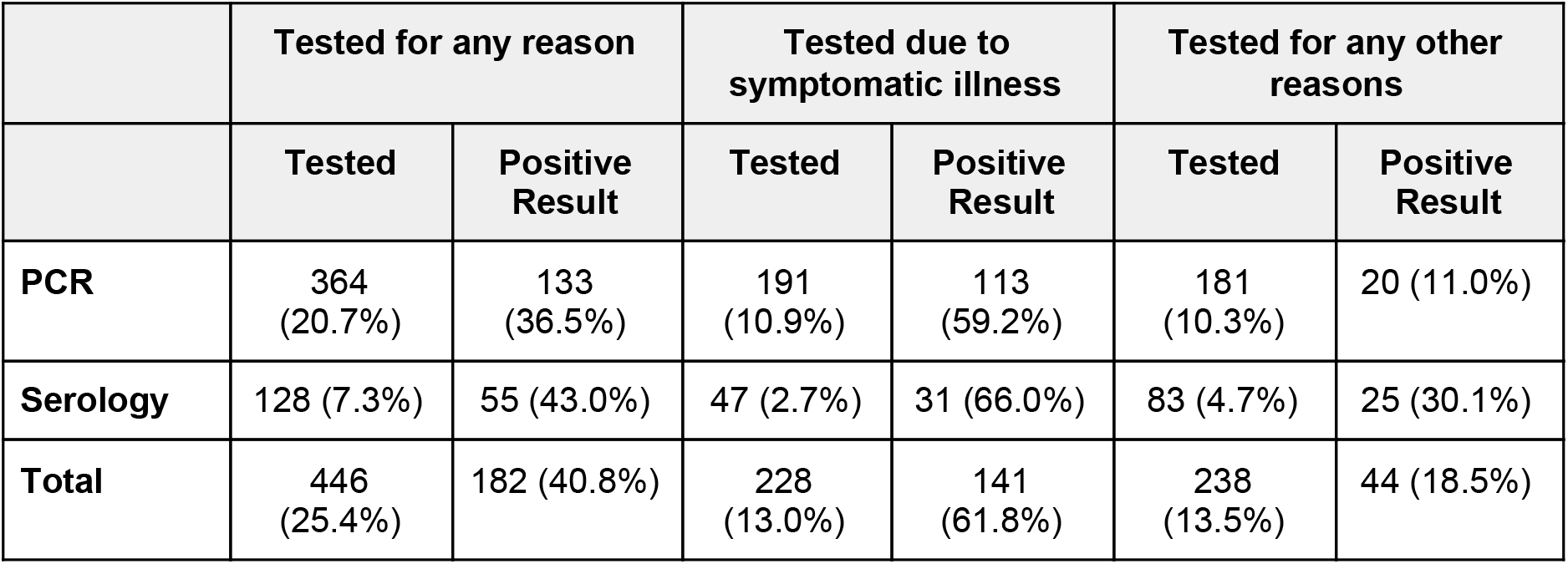
Characteristics of the tests undertaken, stratified by presence of symptoms.

|  | Tested for any reason |  | Tested due to symptomatic illness |  | Tested for any other reasons |  |
| --- | --- | --- | --- | --- | --- | --- |
|  | Tested | Positive Result | Tested | Positive Result | Tested | Positive Result |
| **PCR** | 364<br>(20.7%) | 133<br>(36.5%) | 191<br>(10.9%) | 113<br>(59.2%) | 181<br>(10.3%) | 20 (11.0%) |
| **Serology** | 128 (7.3%) | 55 (43.0%) | 47 (2.7%) | 31 (66.0%) | 83 (4.7%) | 25 (30.1%) |
| **Total** | 446<br>(25.4%) | 182 (40.8%) | 228<br>(13.0%) | 141<br>(61.8%) | 238<br>(13.5%) | 44 (18.5%) |

### Routine Testing for SARS-CoV-2

Amongst randomly selected households, a total of 446 individuals (25.4%) had undertaken either PCR or serological testing for SARS-CoV-2 and 182 individuals (10.3%) reported a positive result on at least one test. Reported reasons for testing were because of symptomatic disease or stated as “for other reasons” (Table 1). Individuals who reported testing were older (median 31 IQR 15-46 vs 12 IQR 5-25) and more likely to be male (52.7% vs 47.1%).

Overall 228 individuals (13%) underwent either PCR and/or serological testing because of symptomatic illness. A total of 191 symptomatic individuals reported providing a sample for PCR with a swab positivity rate of 59% (n = 113). By comparison 20 of 181 PCR tests done for other reasons were positive (11%). Overall 182 individuals (10.3%) reported already having received a positive test for SARS-CoV-2 prior to the survey.

Serology on this cohort revealed an overall seroprevalence for SARS-CoV-2 of 64.3% (95% CI 61.6-67.0%, 799/1,242). The seroprevalence varied by age between 27.6% (95% confidence interval (CI) 20.8 - 35.6%) for children aged under 5 years of age to 73.8% (95% CI 68.2 - 78.8%) amongst secondary school children and 74% (95% CI 70.0 −77.6%) adults (Figure 2) (Supplementary Table 2). Seroprevalence was significantly higher amongst men (68.8%, 95% CI 64.9 - 72.5%) than women (59.7%, 95% CI 55.8 - 63.5%) (*p* = 0.001). Only three individuals (2%) reported a previous positive PCR result but did not have detectable anti-spike antibodies. All three individuals had a positive PCR in October and therefore a negative serological test might reflect that they had not yet seroconverted at the time of sample collection. In a multivariable random-effects logistic regression model, seropositivity was associated with increasing age, male gender, household density and being unable to work from home, but not associated with attendance at community gatherings (Table 2). No pre-existing comorbidities were associated with seropositivity.

**Table 2.** Association between seropositivity and demographic and behavioural variables.*Likelihood ratio test

|  |  | Unadjusted Odds Ratio |  | Adjusted Odds Ratio |  |
| --- | --- | --- | --- | --- | --- |
|  |  | OR | p-value* | aOR (95% CI) | p-value* |
| **Age** | **Young Children (0-4 years)** | 0.07 (0.04 - 0.12) | <0.0001 | 0.10 (0.05-0.20) | <0.0001 |
|  | **Primary Education (5-10 Years)** | 0.36 (0.24-0.54) |  | 0.77 (0.37-1.61) |  |
|  | **Secondary Education (11-18 Years)** | 0.87 (0.58-1.31) |  | 1.70 (0.89-3.27) |  |
|  | **Working age Adults (19-67)** | 1 |  | 1 |  |
|  | **Retirement age adults (67+ Years)** | 0.32 (0.13-0.80) |  | 0.35 (0.14-0.91) |  |
| **Gender** | **Female** | 1 | 0.009 | 1 | 0.002 |
|  | **Male** | 1.46 (1.10 - 1.93) |  | 1.62 (1.18-2.24) |  |
| **Housing** | **Not overcrowded** | 1 | 0.49 | 1 | 0.4 |
|  | **Overcrowded** | 0.86 (0.55 - 1.34) |  | 1.23 (0.74-2.06) |  |
| **Employment and Education Status** | **Neither Working nor in Education** | 1 | <0.001 | 1 | 0.06 |
|  | **In Education** | 0.49 (0.35 - 0.67) |  | 0.45 (0.25-0.81) |  |
|  | **Working From Home** | 1.78 (1.1 -2.87) |  | 1.39 (0.82-2.38) |  |
|  | **Working Outside Home** | 1.25 (0.68-2.3) |  | 0.81 (0.41-1.58) |  |
| **Number of Community Gatherings Attended** |  | 1.02 (0.98-1.06) | 0.34 | 1.04 (0.84-1.28) | 0.7 |

Overall, 697 (37.5%) individuals reported an illness they thought was consistent with COVID-19. There were clear peaks in reported illness consistent with the first and second waves of COVID-19 in the United Kingdom (Figure 3). Of individuals reporting a suspected illness 49.8% were male and the median age was 28 (IQR 14-41). A total of 16 (0.9%) individuals reported hospitalisation for COVID-19 and a further three individuals were reported to have died of COVID-19.

Overall 81.9 % of individuals who reported an illness consistent with COVID-19 were sero-positive and 53.7% of asymptomatic individuals were sero-positive. Of cardinal symptoms of COVID-19, self-reported cough (OR 3.0, 95% CI: 2.1-4.2), fever (OR 2.3, 95% CI: 1.6 −3.4) and loss of smell or taste (OR 11.8, 95% CI: 6.8 −20.6) were all associated with seropositivity. In the overall population self-reported loss of smell or taste had a positive predictive value of 94.5% for positive serology (Supplementary Table 3).

**Table 3.** Association between log-transformed antibody titres and participant characteristics.

|  | Anti-Spike Antibody Titre |  |  | Anti-Receptor Binding Domain Titre |  |  | Anti-Nucleocapsid Antibody Titre |  |  |
| --- | --- | --- | --- | --- | --- | --- | --- | --- | --- |
|  | Beta | 95% CI | p-value | Beta | 95% CI | p-value | Beta | 95% CI | p-value |
| All Patients |  |  |  |  |  |  |  |  |  |
| Male Gender | 0.165 | 0.061-0.270 | 0.002 | 0.181 | 0.084-0.278 | 0.0003 | 0.135 | 0.051-0.220 | 0.002 |
| Age (years) | 0.003 | 0.000-0.006 | 0.095 | 0.003 | 0.000-0.006 | 0.031 | 0.004 | 0.002-0.007 | 0.001 |
| Symptomatic Illness | 0.543 | 0.422-0.665 | <0.001 | 0.447 | 0.335-0.559 | <0.0001 | 0.441 | 0.344-0.539 | <0.0001 |
| Symptomatic Individuals |  |  |  |  |  |  |  |  |  |
| Male Gender | 0.057 | -0.09 - 0.206 | 0.448 | 0.061 | -0.075 - 0.198 | 0.381 | 0.033 | -0.094 - 0.160 | 0.612 |
| Age (years) | 0.010 | 0.006 - 0.014 | <0.0001 | 0.009 | 0.005 - 0.013 | <0.0001 | 1.23 | 0.009 - 0.016 | <0.0001 |
| Time since reported illness (days) | -0.001 | -0.00004 - -0.002 | 0.041 | -0.001 | 0.000 - 0.002 | 0.022 | -0.002 | -0.001 - -0.003 | <0.0001 |

Titres of spike, receptor binding domain and nucleocapsid antibodies were higher amongst individuals who reported a symptomatic illness (Figure 4) (Table 3). Amongst symptomatic individuals titres declined following time since reported symptomatic illness which was more marked for nucleocapsid than for other targets (Supplementary Figure 1).

**Figure 4.**
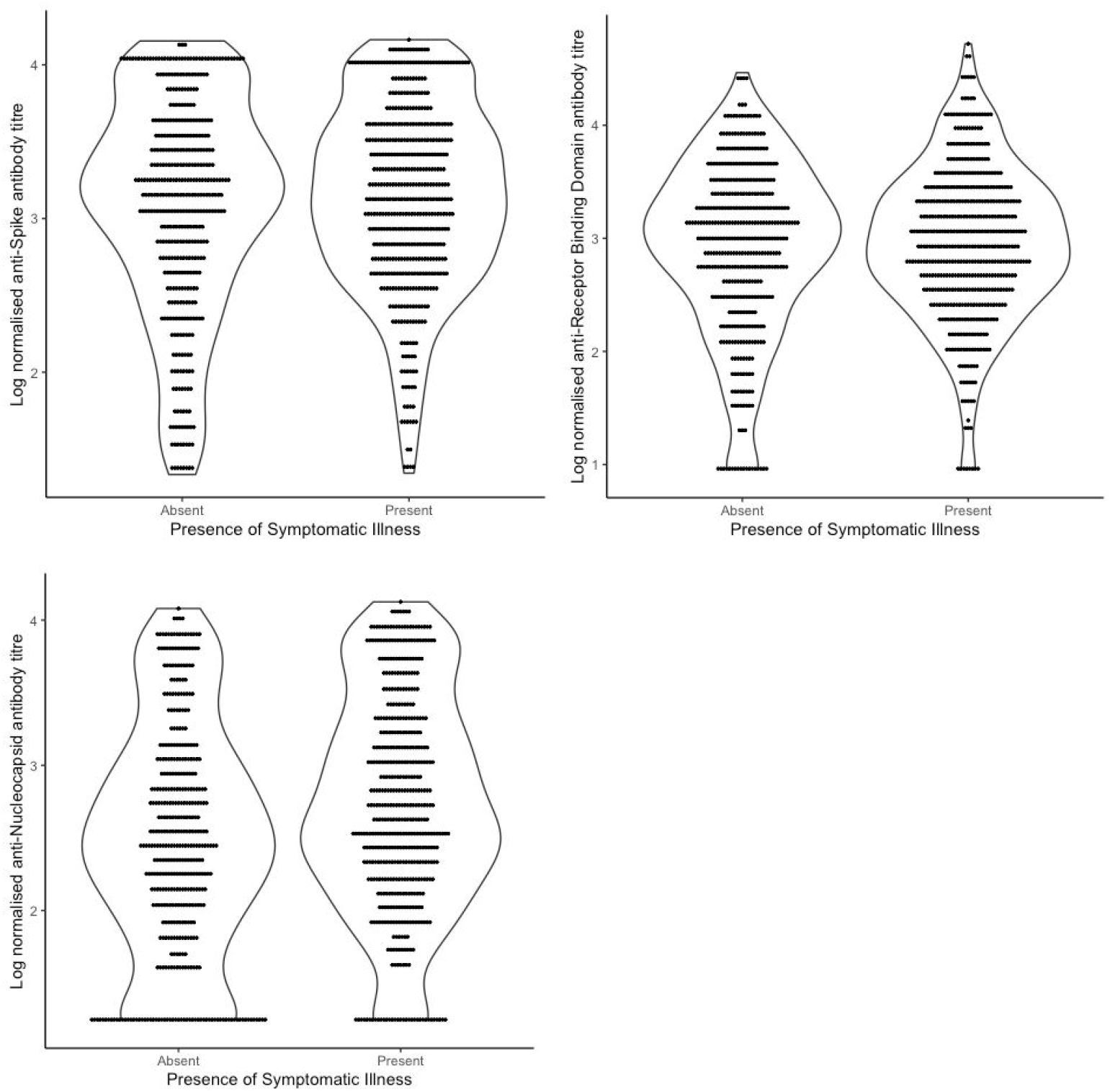
Log normalised antibody titres for anti-Spike, anti-Receptor Binding Domain and anti-Nucleocapsid. Each panel is stratified by reporting of COVID-19-like symptoms by the participant. The shape shows the density of the distribution of samples.

## Discussion

We found an extremely high seroprevalence of SARS-CoV-2 antibodies in a strictly Orthodox Jewish community in the UK. Our estimate of 65% population seroprevalence is markedly higher than recent estimates of 6.9% (95%CI 6.3-7.4%) nationally and 10.8% (95% CI 9.3-12.5%) in London by random sampling in October by the Office for National Statistics (ONS)^20^. Rapid declines in self-reported illness followed the introduction and adherence to lockdown in March, demonstrating that even in this highly connected community such measures are effective at reducing transmission. However over the course of 2020, the overall seroprevalence in this tightly knit religious community reached levels similar to those seen in Manaus, Brazil where a seroprevalence of more than 65% has been reported in adults^21^. As our survey was completed by early December 2020, prior to the subsequent winter case surge in London, it is likely that the overall burden of infection in this community is now even higher.

Our estimates are amongst the highest sero-prevalence of SARS-CoV-2 described anywhere in the world to date. The precise reasons why the burden of SARS-CoV-2 has been so high in this population are unclear. In Israel, Orthodox Jewish communities had markedly higher SARS-CoV-2 PCR swab positive incidence rates compared to other socio-economically similar communities during the first wave of the Israeli epidemic^14,15^. Data from other sources suggest that lower socio-economic status, ongoing need to travel to work and a greater burden of pre-existing comorbidities, may all contribute to increased risk of acquiring SARS-CoV-2.

In our study population there was high seroprevalence in all age groups, with the highest seroprevalence in working-age adults and older children where seroprevalence reached 74%. A strength of our study is the extremely high number of children recruited, which reflects the high total fertility rates amongst strictly Orthodox Jewish women^12,13^. Seroprevalence in the youngest children was lower at 27% but rose rapidly to more than 50% amongst primary school-aged children. The seroprevalence found in this group is approximately four times that reported in a UK multicentre study on healthcare worker children aged 2-15 years demonstrating the high rates of infection in all age groups in the current study ^22^.

Seroprevalence was higher amongst men than women in our study. Higher SARS-CoV-2 attack rates in men compared to women have also been reported in other contexts^23^. The higher rate of infection amongst men might reflect biological differences, differences in comorbidities, differences in social mixing patterns or other behaviours. We did not find any significant associations with reported attendance at community events, workplace or household overcrowding in this population which may be because of a true absence of effect, or that due to the extremely high seroprevalence, the ability to detect risk factors is limited.

The majority of reported symptomatic illness occurred during the first and second waves of the COVID-19 pandemic mirroring case reporting patterns seen across London and the UK. Just over 10% of participants reported at least one swab test due to symptomatic illness since March, with an overall swab positivity rate of 59%. In line with other studies, children were significantly less likely to report a symptomatic illness^24,25^.

We found that antibody titres against spike and nucleocapsid proteins declined at different rates, and in line with other studies^16,26^, that anti-nucleocapsid declined more quickly. If we had used only anti-nucleocapsid antibodies as a marker of previous infection our estimated seroprevalence would have been 42.8%, significantly under-estimating the likely size of the epidemic that has occurred in this population. Declines in anti-spike antibody titre were less marked over time but it is still possible that we have under-estimated the true seroprevalence in this population. Titres of both spike and nucleocapsid antibodies were higher amongst individuals who reported symptomatic illness.

Our study has a number of limitations. We recruited 38% of households that were approached and obtained serum samples from 70% of study participants. This enrollment rate is similar to other national COVID-19 household surveillance studies, such as the ONS COVD-19 Infection survey^27^, suggesting it is unlikely to be a major source of bias. Individuals who gave serum samples were slightly older than those from whom serum was unavailable which may result in an over-estimation of the overall population seroprevalence. We relied on self-report of presumed COVID-19 illness which may be unreliable. However the timings of self reported illness match well to national surveillance data and self-reported illness was strongly associated with the presence of anti-spike antibodies suggesting that in this population this was a reliable metric.

Although our study was conducted in a strictly Orthodox Jewish community, these communities share many characteristics with other ethnic and religious minority groups including larger family sizes, increased population density, children attending select schools, regular attendance at communal events and gatherings, and English as a second language. As such our findings are likely relevant to other tightly-knit ethnic and religious minority groups in the UK and elsewhere. Our work, conducted in direct collaboration with the community, should be a model for understanding risk in minority populations where there are no similar published data currently.

In conclusion, we found evidence for an extremely high rate of SARS-CoV-2 infection in this specific community affecting individuals of all ages. This provides further evidence that minority communities in the UK and elsewhere are disproportionately affected by the COVID-19 pandemic. The reasons for this remain unclear, although are likely to be a complex interplay of socio-economic and behavioural factors. Further studies to better understand drivers of transmission in ethnic and religious minority populations, conducted wherever possible in partnership with communities themselves, are urgently needed to reduce health disparities and improve outcomes for these populations.

## Data Availability

A de-identified data set with the study protocol will be available on request by email.

## Supplementary Data

1. Supplementary Appendix
2. STROBE Checklist

## Contributors

MM, RME, ChR, DG conceived of the study. KG and DL co-ordinated the survey. KG, MJ, VG conducted lab work. SL, WW, BK, TC, NS contributed to the design of the study. MM, KG, WW, RME verified the underlying data. All authors contributed to the analysis and writing of the manuscript.

## Data sharing

Data available on request

## Acknowledgements

We thank Elizabeth Miller for helpful discussions on study design, and Daniel Grint for statistical queries. RME reports funding from HDR UK (grant: MR/S003975/1), MRC (grant: MC_PC 19065), NIHR (grant: NIHR200908). MM reports funding from UKRI and NIHR (application COV0335, grant: MR/V027956/1). KG reports funding from the Wellcome Trust (grant: 210830/Z/18/Z)

