## Supplementary files for "Extremely high SARS-CoV-2 seroprevalence in a strictly-Orthodox Jewish community in the UK"

#### 1. Demographics

**Supplementary Table 1: Survey respondent demographics**

| **Variable** | | **Frequency** |
|---|---|---|
| **Gender** | Male | 853 (48.5%) |
| | Female | 906 (51.5%) |
| **Age** | Median (IQR) | 14 years (7-33) |
| **Age Group** | Early Years (0-4 years) | 307 (17.5%) |
| | Primary School (5-10 years) | 357 (20.3%) |
| | Secondary School (11-18 years) | 360 (20.5%) |
| | Adults (19-66 years) | 684 (38.9%) |
| | Retirement Age Adults (67+) | 51 (2.9%) |
| **Education and Employment** | In formal education | 776 (44.1%) |
| | Working from home | 238 (13.5%) |
| | Working outside home | 133 (7.6%) |
| | Neither in education or formal employment | 612 (34.8%) |
| **Self Reported Comorbidities** | Asthma | 11 (0.6%) |
| | COPD | 2 (0.1%) |
| | Hypertension | 31 (1.8%) |
| | Diabetes | 21 (1.2%) |
| | Cardiovascular Disease | 9 (0.5%) |
| | Chronic Kidney Disease | 1 (0.1%) |
| | Dementia | 0 (0%) |

### 2. Antibody Titres and time since self-reported COVID-19-like symptoms

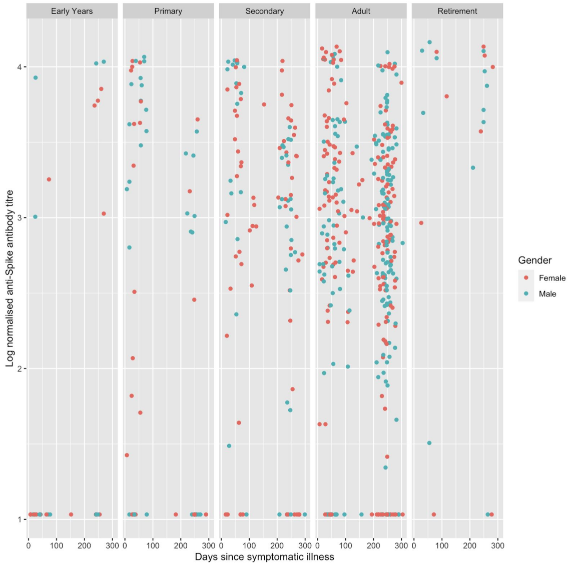


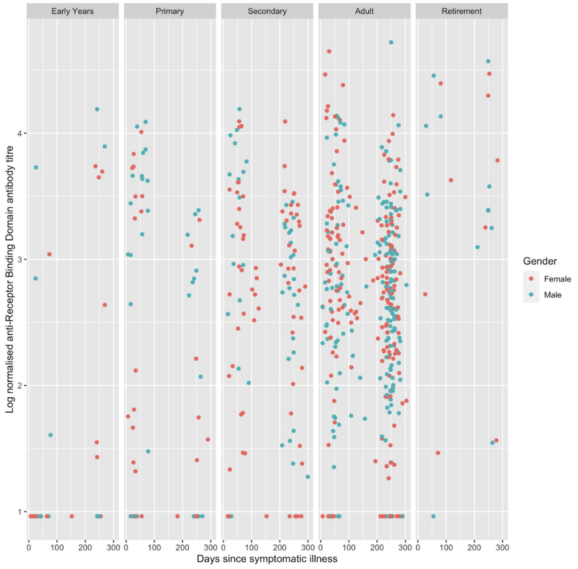

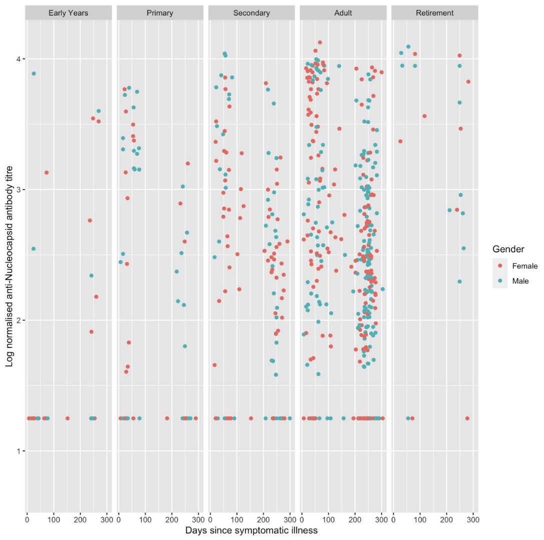


**Supplementary Figure 1. Log normalised antibody titres against spike, receptor binding domain and nucleocapsid antigens by time since self-reported COVID-19 symptoms.** Values are shown stratified by age (panels) and gender (colour).

3. **Antibody Seroprevalence by antibody target and age group**

**Supplementary Table 2: Age stratified seroprevalence.**

| Age Group | anti-Spike SARS-CoV-2 antibodies | anti-Receptor Binding Domain antibodies | anti-Nucleocapsid SARS-CoV-2 antibodies |
|---|---|---|---|
| Early Years (0-4 years) | 27.6% (20.8-35.6%) | 22.4% (16.2 - 30.0%) | 18.4% (12.8 - 25.7%) |
| Primary School (5-10 years) | 56.4% (49.8-62.7%) | 43.8% (41.8 - 54.9%) | 42.8% (36.4-49.4%) |
| Secondary School (11-18 years) | 73.8% (68.2-78.8%) | 65.6% (59.7 - 71.1%) | 50.9% (44.9-56.9%) |
| Adults (19-66 years) | 74% (70.0-77.6%) | 57.8% (53.5-62.0%) | 45.4% (41.1-49.7%) |
| Retirement Age Adults (67+) | 54.8 (38.8-69.8%) | 40.5% (26.0-56.7%) | 45.2% (30.2-61.2%) |

4. **Positive and negative predictive value of symptoms by age group**

**Supplementary Table 2: Positive and negative predictive values stratified by age for symptoms reported as COVID-19-like illness.** PPV = positive predictive value, NPV = negative predictive value.

| | Fever | | Cough | | Loss of Smell or Taste | |
|---|---|---|---|---|---|---|
| **Age Group** | **PPV** | **NPV** | **PPV** | **NPV** | **PPV** | **NPV** |
| Overall | 79.0% | 38.3% | 81.6% | 43.6% | 94.1% | 42.7% |
| Early Years (0-4 years) | 33.3% | 68.2% | 36.4% | 76.1% | 33.3% | 68.1% |
| Primary School (5-10 years) | 57.1% | 42.9% | 65.2% | 48.3% | 100% | 44.6% |
| Secondary School (11-18 years) | 63.0% | 25.0% | 73.0% | 28.0% | 88.9% | 29.1% |
| Adults (19-66 years) | 90.9% | 31.0% | 91.4% | 35.9% | 96.6% | 37.1% |
| Retirement Age Adults (67+) | 77.8% | 51.5% | 90.0% | 56.3% | 80.0% | 48.6% |

5. **Sensitivity Analysis**

In a sensitivity analysis in which the threshold for spike positivity was doubled, seroprevalence was 49.7%.

6. **Population Structure**

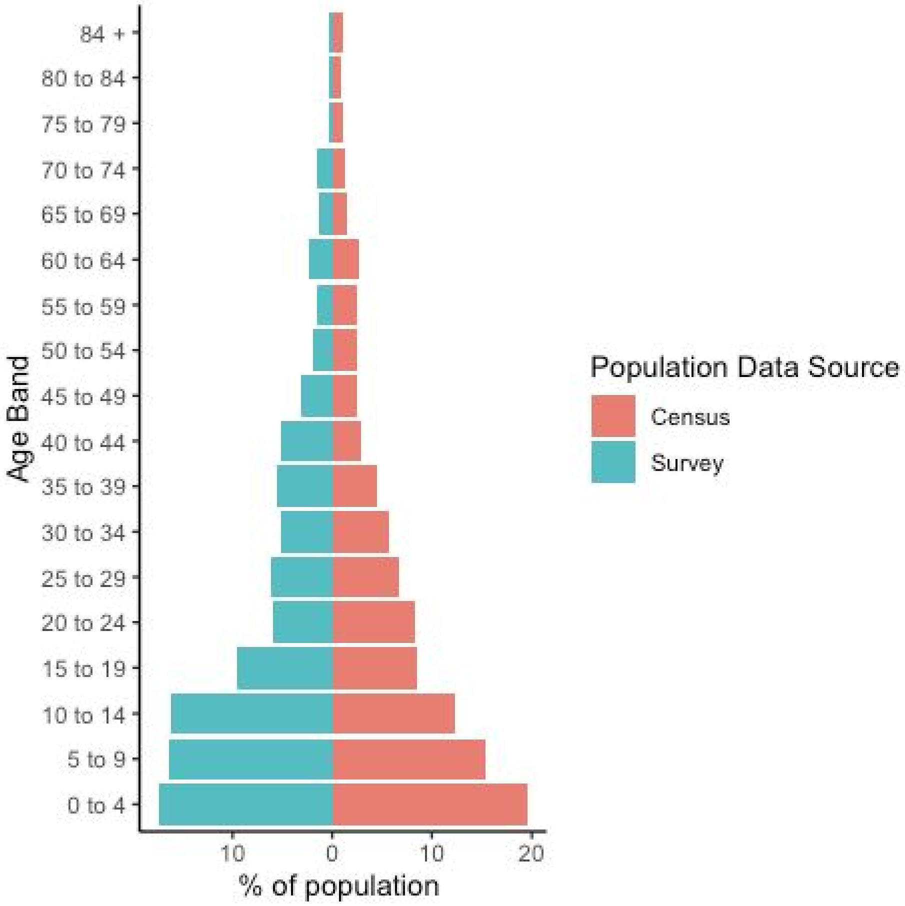


**Supplementary Figure 2. Population age structure for survey respondents compared to the overall Haredi population.** Census data is from 2011.

7. **Consort Diagram**

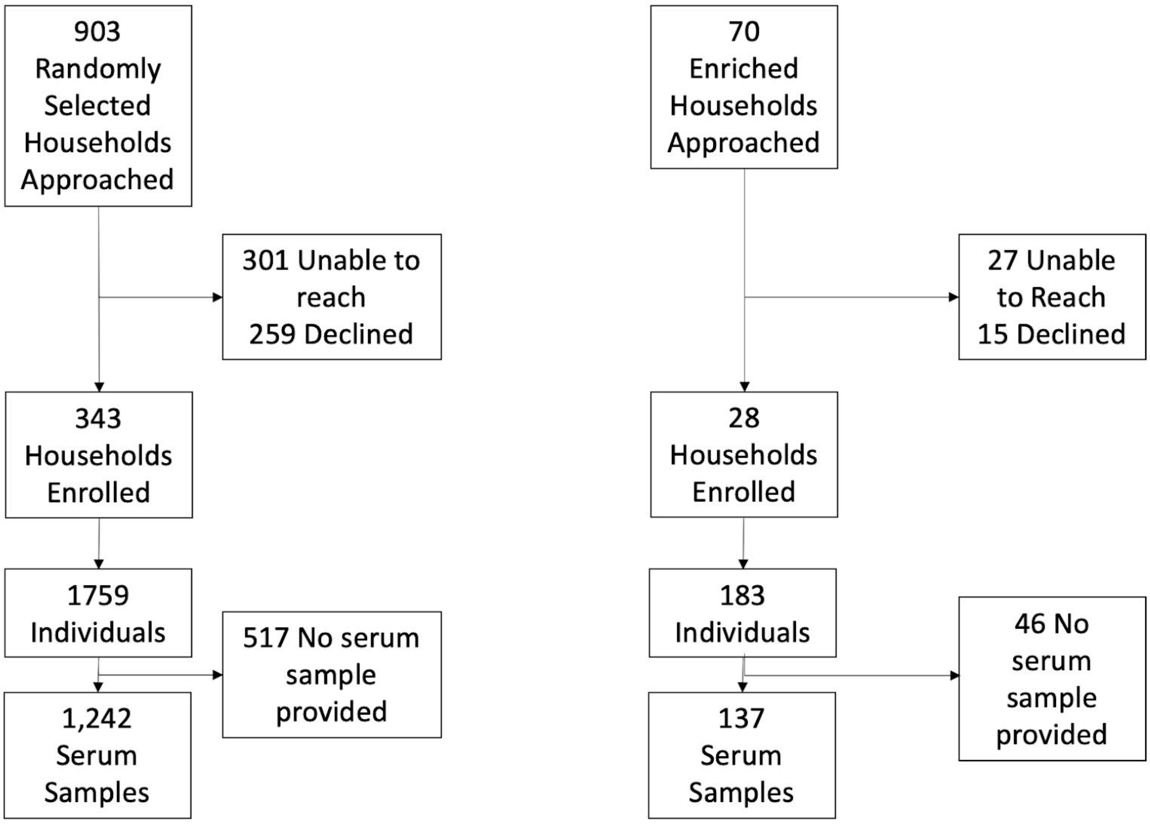


**Supplementary Figure 3. Consort diagram showing enrollment of both the randomly selected and the enriched households into the study**
